# *APOE* loss-of-function variants: Compatible with longevity and associated with resistance to Alzheimer’s Disease pathology

**DOI:** 10.1101/2023.07.20.23292771

**Authors:** Augustine Chemparathy, Yann Le Guen, Sunny Chen, Eun-Gyung Lee, Lesley Leong, John Gorzynski, Guangxue Xu, Michael Belloy, Nandita Kasireddy, Andrés Peña Tauber, Kennedy Williams, Ilaria Stewart, Thomas Wingo, James Lah, Suman Jayadev, Chad Hales, Elaine Peskind, Daniel D Child, C Dirk Keene, Le Cong, Euan Ashley, Chang-En Yu, Michael D. Greicius

**Affiliations:** Department of Neurology and Neurological Sciences, Stanford University School of Medicine, Stanford, CA; Quantitative Sciences Unit, Department of Medicine, Stanford University School of Medicine, Stanford, CA; Geriatric Research, Education, and Clinical Center, VA Puget Sound Health Care System, Seattle, WA; Department of Genetics, Stanford University School of Medicine, Stanford, CA; Department of Pathology, Stanford University School of Medicine, Stanford, CA; Emory University School of Medicine, Atlanta, GA; Goizueta Alzheimer’s Disease Center, Emory University School of Medicine, Atlanta, GA; Department of Neurology, Emory University School of Medicine, Atlanta, GA; Department of Neurology, University of Washington, Seattle, WA; Veterans Affairs Northwest Network Mental Illness Research, Education, and Clinical Center, Veteran Affairs Puget Sound Health Care System, Seattle, WA; Department of Psychiatry and Behavioral Sciences, University of Washington, Seattle, WA; Department of Laboratory Medicine and Pathology, University of Washington School of Medicine, Seattle, WA; Center for Inherited Cardiovascular Disease, Stanford University, Stanford, CA; Division of Cardiology, Department of Medicine, Stanford University School of Medicine, Stanford, CA; Department of Medicine, University of Washington, Seattle, WA

**Keywords:** Alzheimer’s Disease, Neurodegenerative disorders, Human genetics, Loss-of-function, Apolipoprotein E

## Abstract

The ε4 allele of apolipoprotein E (*APOE*) is the strongest genetic risk factor for sporadic Alzheimer’s Disease (AD). Knockdown of this allele may provide a therapeutic strategy for AD, but the effect of *APOE* loss-of-function (LoF) on AD pathogenesis is unknown. We searched for *APOE* LoF variants in a large cohort of older controls and patients with AD and identified six heterozygote carriers of *APOE* LoF variants. Five carriers were controls (ages 71-90) and one was an AD case with an unremarkable age-at-onset between 75-79. Two *APOE* ε3/ε4 controls (Subjects 1 and 2) carried a stop-gain affecting the ε4 allele. Subject 1 was cognitively normal at 90+ and had no neuritic plaques at autopsy. Subject 2 was cognitively healthy within the age range 75-79 and underwent lumbar puncture at between ages 75-79 with normal levels of amyloid. The results provide the strongest human genetics evidence yet available suggesting that ε4 drives AD risk through a gain of abnormal function and support knockdown of *APOE* ε4 or its protein product as a viable therapeutic option.

## Introduction

Advancements in genetic engineering have resulted in treatments for monogenic neurological disorders previously considered intractable. The most tangible progress has occurred in spinal muscular atrophy (SMA), a neurodegenerative disease caused by the loss-of-function of *SMN1*. The antisense oligonucleotide (ASO) nusinersen increases the attainment of motor milestones in infants with SMA by altering splicing of the paralogous *SMN2* gene to rescue its function. Risdiplam, a small molecule acting on *SMN2*, and abeparvovec, an AAV-based therapy that restores *SMN1,* have also received FDA approval^1^. Knockdown approaches using ASOs have been employed in familial amyotrophic lateral sclerosis and Huntington’s disease^2, 3^. While these ASO trials did not result in clinical efficacy, they significantly reduced the abnormal protein in both disorders and informed new trials. These gene-targeting therapies offer hope that similar approaches could be successful in Alzheimer’s disease (AD).

Individuals carrying the ε4 allele of apolipoprotein E (*APOE*) have a significantly elevated risk of AD, suggesting that genetic modulation of *APOE* ε4 could be therapeutic. An *APOE* ε4/ε4 individual of European ancestry has a sixteen-fold increase in AD risk and eighteen year earlier age at AD onset compared to a European ancestry individual carrying two copies of the more prevalent *APOE* ε3 allele^4^. A critical question regarding pathogenesis is whether ε4 is inherently detrimental (in which case one would want to knock it down) or is less functional than ε3 (in which case one might want to increase levels of the protein in ε4 homozygotes). Evidence from animal models of AD can be marshaled to support either possibility^5, 6^. The bulk of the animal literature supports knockdown of *APOE* as likely to reduce AD pathogenesis. For example, studies have shown that reducing *APOE* results in reduced amyloid^6–8^ and tau pathology^9^ in animal models. However, other animal studies support increasing *APOE* as a potential therapeutic approach^10–12^.

Evidence from human studies is generally lacking because *APOE* loss-of-function (LoF) variants are rare. Only one individual carrying an *APOE* LoF variant has been cognitively assessed and reported in the literature^13^. The patient was a man in the age range 40-44 homozygous for *APOE* c.291del (p.E97fs) and presenting with severe hyperlipidemia. He had negative spinal fluid biomarkers for AD but impaired memory^14^. Despite the normal biomarkers, the cognitive impairment and relatively young age make this case ultimately uninformative on the question of whether reducing apoE might be beneficial or detrimental in terms of AD pathogenesis. We have found no publications describing AD-relevant phenotypes in older subjects heterozygous for *APOE* LoF variants. Here we ask whether *APOE* LoF variants impact AD pathogenesis.

## Results

To characterize AD phenotypes of *APOE* LoF individuals, we searched the Alzheimer’s Disease Sequencing Project (ADSP) whole-exome and whole-genome sequencing datasets (**Table 1**) for predicted *APOE* LoF single nucleotide polymorphisms (SNPs) and structural variants (SVs). All SNPs on *APOE* were extracted, annotated with predicted variant type, and filtered for predicted high impact variants affecting the canonical transcript (**Supplementary Table 1**). The most common end-truncation variant, rs121918396 (p.W228*), was expressed in human hepatocytes and shown not to affect apoE protein level (**Figure S1**); thus predicted end-truncation variants were not explored further. We identified five participants carrying *APOE* LoF SNPs – three carriers of rs777551553 (p.W5*), one carrier of rs923895447 (p.L8*) and one carrier of 19:44907831:C:T (p.Q39*). These five carriers were normal at their last cognitive assessment (mean age at last assessment = 82, range 71-90). We also searched the ADSP SV dataset for insertions, deletions, duplications, inversions, and translocations overlapping *APOE* and further filtered these SVs for those predicted to be LoFs (**Supplementary Table 1**). We identified one individual with AD carrying a 1,798 base pair deletion (19:44905303-44907102) that eliminates the bulk of the *APOE* promoter region and exons 1 and 2 (including the start codon and the signal peptide). *APOE* genotype, age, and diagnoses of the six *APOE* LoF variant carriers are shown in **Figure 1a** with variant positions shown in **Figure 1b**.

**Figure 1.**
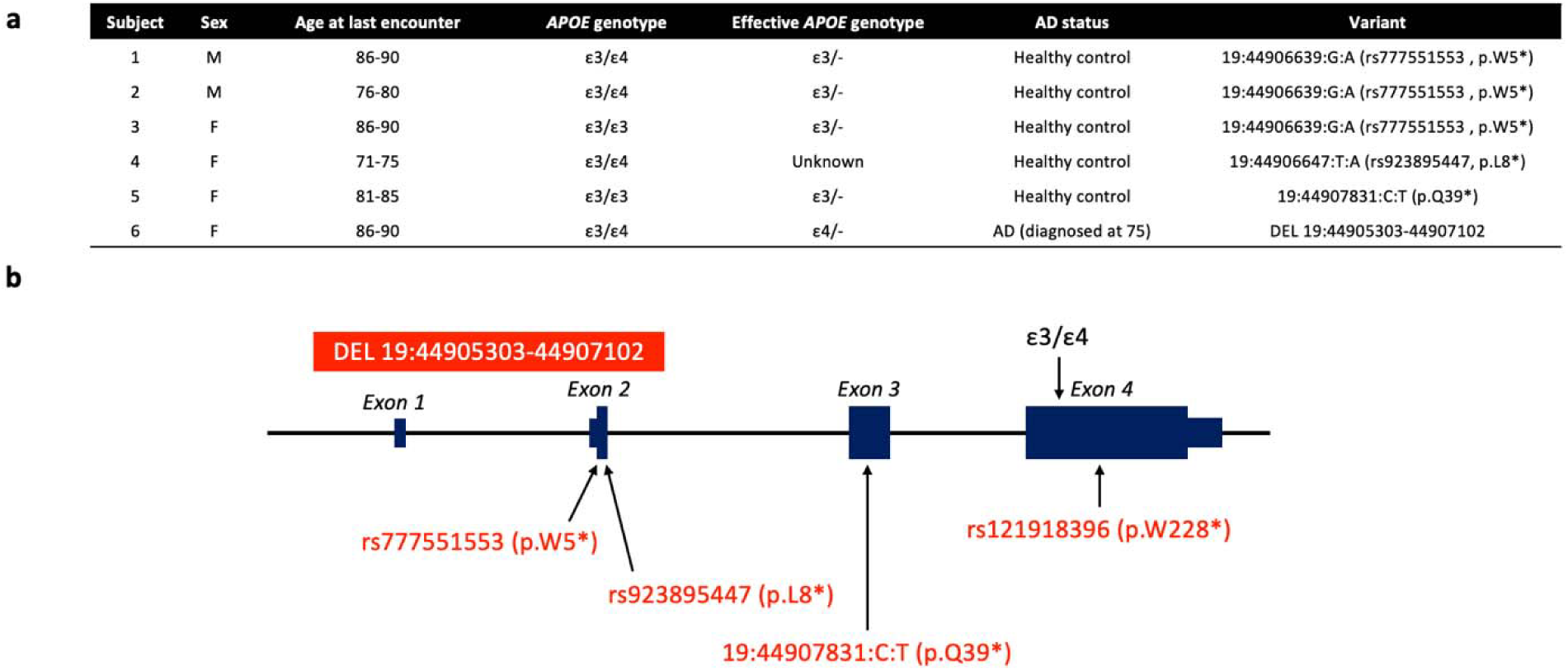
*APOE* loss-of-function carrier demographics and variant positions. (a) Carrier demographics. Six carriers of high confidence *APOE* loss-of-function variants were identified among 26,605 older controls and 20,856 AD cases sequenced as part of the Alzheimer’s Disease Sequencing Project. (b) Three distinct single nucleotide polymorphisms and one structural variant were identified. Genomic coordinates are based on hg38.

**Table 1:** Demographics of ADSP cohorts. Abbreviations: Whole-exome sequencing (WES), Whole-genome sequencing (WGS), Diagnosis (Dx), Alzheimer’s Disease (AD), Control (CN)

| Cohort | Dx | N | Sex | Age | APOE |  |  |  |  |  |
| --- | --- | --- | --- | --- | --- | --- | --- | --- | --- | --- |
|  |  |  | Female (%) | Median age [IQR] | 22<br>N (%) | 23<br>N (%) | 33<br>N (%) | 34<br>N (%) | 44<br>N (%) | 24<br>N (%) |
| ADSP WES | AD | 8723 | 61.8% | 75[70-82] | 29(0.3%) | 540(6.2%) | 3884(44.5%) | 3462(39.7%) | 568(6.5%) | 237(2.7%) |
|  | CN | 9617 | 63.4% | 84[76-89] | 83(0.9%) | 1530(15.9%) | 5762(59.9%) | 1864(19.4%) | 133(1.4%) | 244(2.5%) |
|  | Other | 2163 | 51.4% | 75[69-82] | 17(1.0%) | 209(11.8%) | 984(55.7%) | 455(25.8%) | 60(3.4%) | 41(2.3%) |
|  | Total | 20503 | 61.5% | 79[72-87] | 129(0.6%) | 2279(11.3%) | 10630(52.9%) | 5781(28.8%) | 761(3.8%) | 522(2.6%) |
| ADSP WGS | AD | 12133 | 60.4% | 72[64-80] | 27(0.2%) | 558(4.6%) | 5003(41.2%) | 4943(40.7%) | 1327(10.9%) | 270(2.2%) |
|  | CN | 16988 | 63.7% | 73[66-81] | 78(0.5%) | 1605(9.4%) | 10735(63.2%) | 3900(23.0%) | 357(2.1%) | 311(1.8%) |
|  | Other | 7240 | 55.8% | 75[69-81] | 33(0.5%) | 645(8.9%) | 4111(57.0%) | 1953(27.1%) | 316(4.4%) | 151(2.1%) |
|  | Total | 36361 | 61.0% | 73[65-80] | 138(0.38%) | 2808(7.7%) | 19849(54.6%) | 10796(29.7%) | 2000(5.5%) | 732(2.0%) |

Subject 1 (carrier of rs777551553_A (p.W5*)) demonstrated striking resistance to amyloid pathology given his age at death (age 90+) and ε3/ε4 genotype. While he had evidence of hyperphosphorylated tau pathology (Braak stage IV of VI) there was no appreciable amyloid β (Aβ) pathology in the brain (CERAD neuritic plaque score 0 of 3, Thal Aβ stage 0 of 5^15^, and no cerebral amyloid angiopathy). The absence of amyloid pathology and moderate extent of tau pathology at advanced age make this individual an outlier among ε3/ε4 heterozygotes (**Figure 2**). rs777551553_A was heterozygous and in phase with ε4 in this individual (**Figure S2a, b**). Sanger sequencing of reverse-transcribed mRNA detected both ε3 and ε4 transcripts (**Figure S2c**), as expected of a stop-gain variant. This variant would prematurely terminate translation to create a truncated 5 amino acid peptide, resulting in an effective ε3/- genotype.

**Figure 2.**
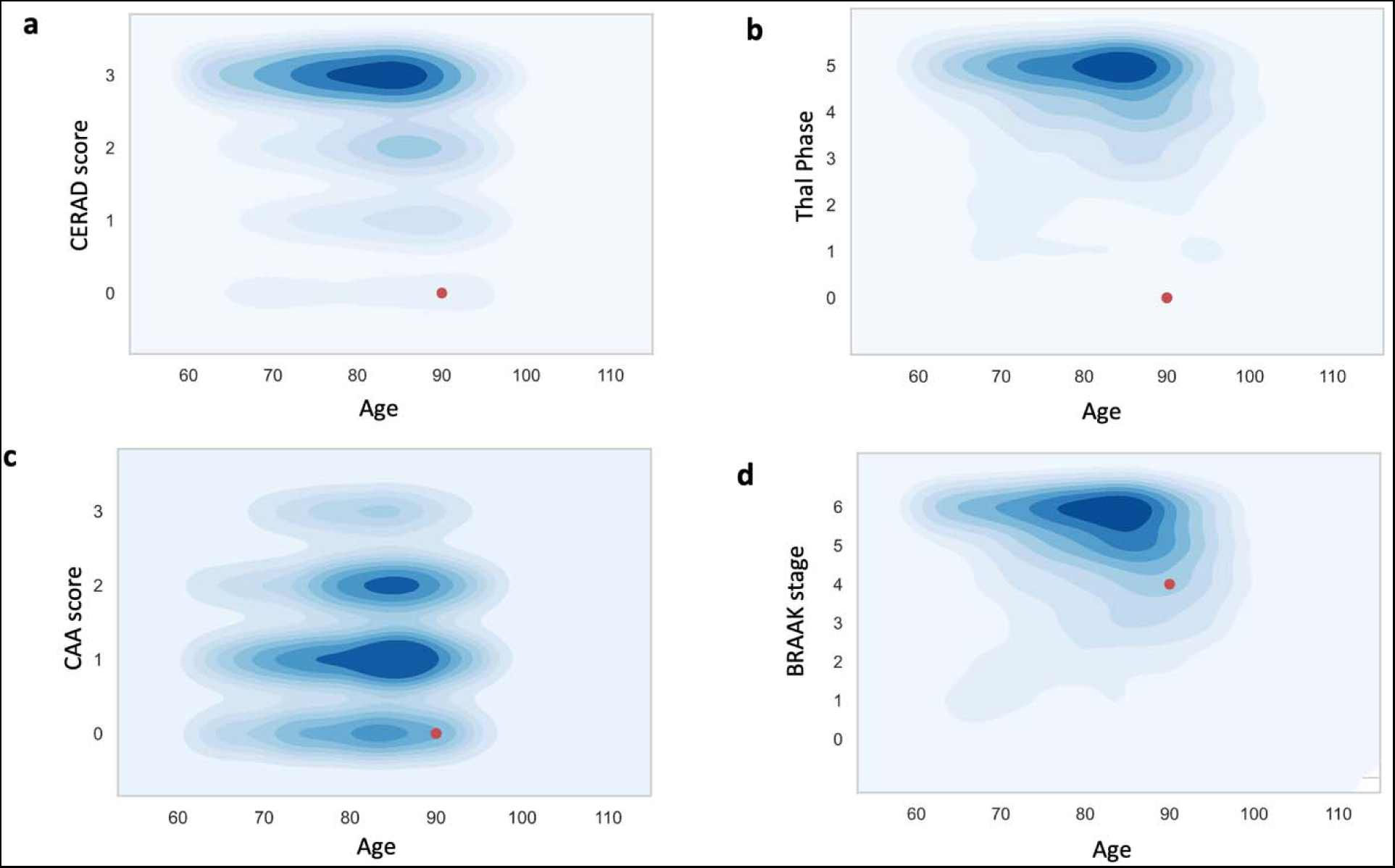
Loss of *APOE* ε4 is associated with absent amyloid pathology and reduced tau pathology in a 90+ year-old control. Subject 1 is a neuropathological outlier among age-matched ε3/ε4 individuals in their (a) CERAD staging of amyloid plaque density; (b) Thal staging of amyloid plaque regional distribution; (c) Cerebral amyloid angiopathy staging; and (d) Braak staging of neurofibrillary tangles (tau pathology).

Subject 2 (also an ε3/ε4 carrier of rs777551553_A (p.W5*)), was cognitively normal at 79. Whole-genome long-read sequencing established that rs777551553_A was in phase with ε4 (**Figure S3**). This subject underwent lumbar puncture at age 75-79 and had normal levels of Aβ and tau. In a meta-analysis of amyloid biomarker studies, by age 75 roughly 2/3rds of ε3/ε4 controls were amyloid-positive by spinal fluid measurement (*Supplementary Figure 3B* in Jansen et al.^16^). This underestimates the “protected” status of Subject 2 because it does not account for the fact that many ε3/ε4 individuals either have mild cognitive impairment or AD by age 75.

Three other carriers of early *APOE* stop-gain mutations were cognitively normal. Subject 3 is a 85-89 year-old ε3/ε3 female, Subject 4 is a 70-74-year-old ε3/ε4 female, and Subject 5 is an 80-84-year-old ε3/ε3 female. Subject 4 did not have mRNA or DNA available for phasing.

Subject 6 is an ε3/ε4 AD patient with a deletion including the *APOE* promoter, start codon, and two of four exons. PCR and Sanger sequencing of the patient’s post-mortem frontal cortex tissue confirmed the deletion (**Figure S4a**) and established that the deletion was heterozygous and in phase with ε3 (**Figure S4b, c**). Only the ε4 allele was detected on Sanger sequencing of reverse transcribed mRNA (**Figure S4d**), indicating that this deletion abolishes transcription of ε3.

Neuropathology in Subject 6 was consistent with AD (CERAD score = 2, Braak Stage VI). This individual had symptom onset at age 75-79, was diagnosed within age range 75-79 and died between ages 85-89. Mean age of onset is 69.73 for ε4 homozygotes (N=1,689) and 73.55 for ε3/4 heterozygotes (N=6,223) in the Alzheimer’s Disease Genetics Consortium (ADGC) and ADSP. Although this patient exclusively expressed the ε4 allele, their disease onset was later than that of a typical ε4/ε4 homozygote suggesting that an ε4/- individual has a preferable disease course to an ε4/ε4 individual. Thus, partial knockdown of ε4 in ε4/ε4 patients may improve the trajectory of AD.

## Discussion

The cognitive phenotypes of *APOE* LoF carriers support the hypothesis that *APOE* ε4 increases risk through a gain of function. The variants identified in this study result in the following *APOE* loss of function mechanisms: stop-gain in the 18 amino acid *APOE* signal peptide that would preclude further translation of apoE (p.W5*, p.L8*, Subjects 1, 2, 3, 4); stop-gain early in the *APOE* coding region that would result in a severely truncated peptide without apoE’s key binding regions (p.Q39*, Subject 5); and deletion involving the *APOE* promoter and exons 1 and 2 (including the start codon and signal peptide) that abolishes APOE transcription altogether (19:44905303-44907102, Subject 6). If *APOE* ε4 increases risk owing to diminished protein function or availability, the *APOE* LoF variants detailed here should be associated with increased risk of AD. Instead, of the six subjects reported here, five were older controls ranging in age from 71-90. The sole AD case had loss of ε3 resulting in an effective ε4/- genotype. If *APOE* ε4 increased risk due to diminished function we would expect such an ε4/- genotype to result in comparable or even increased risk compared to ε4/ε4 homozygotes. Instead, this patient had a later age-at-onset similar to patients with the ε3/ε4 genotype. The phenotypes of these carriers are most consistent with a model in which loss of *APOE* ε4 enhances AD resistance, suggesting that *APOE* ε4 increases AD risk through a gain-of-function.

Most compellingly, we describe two older controls with LoF variants in-phase with ε4 resulting in an effective ε3/- genotype. Subject 1 was cognitively normal at 90 and had no appreciable amyloid pathology. When compared to neuropathological profiles of age- and genotype-matched peers this subject is a clear outlier (**Figure 2**). Subject 2 was cognitively normal within age range 75-79 and had a normal spinal fluid biomarker profile within this age range (seen in only 1/3 of APOE ε3/ε4 controls over 75).

Taken together, these findings support reducing apoE as a therapeutic option in AD. Regarding safety, the identification of 6 long-lived individuals with heterozygous APOE LoF variants suggests that partial knockdown of APOE should be well-tolerated. The resistant phenotypes of two *APOE* ε3/ε4 individuals with a LoF variant on their ε4 allele provides the first human genetics data suggesting that knocking down *APOE* ε4 could reduce AD pathogenesis.

## Author contributions

Conceptualization, data curation, methodology, investigation, writing and revising manuscript: AC, YLG. Methodology, investigation, revising manuscript: SC, EL, LL, JG, GX, MB, NK, APT, KW, IS, DDC. Supervision, revising manuscript: TW, JL, SJ, CH, EP, CDK, LC, EU. Conceptualization, methodology, writing and revising manuscript, supervision: CY, MDG.

## Supplementary material

Supplementary Figures 1-4

Supplementary Table 1: Carriers of *APOE* LoF variant candidates

Supplementary Table 2: Primer designs for characterization of Subjects 1 and 6

## Data sharing statement

Data from the Alzheimer’s Disease Sequencing Project is available via NIAGADS DSS (https://dss.niagads.org/datasets/ng00067).

## Data Availability

Data will be made available after the paper is accepted at a peer-reviewed journal.

## Online Materials and Methods

### Single nucleotide polymorphism identification

36,361 whole-genomes were downloaded from NIAGADS 10^th^ release, 20,504 whole-exomes were downloaded from NIAGADS 4^th^ release (https://dss.niagads.org/datasets/ng00067/#data-releases) and Plink 1.9^17^ was used to extract all SNPs in the range chr19:44905796-44909393 corresponding to *APOE*. ENSEMBL Variant Effect Predictor (VEP)^18^ was used to annotate all SNPs with variant consequence and LoF flags^19^. We filtered for SNPs annotated as high impact (transcript ablation, splice acceptor variants, splice donor variants, stop gains, frameshift variants, stop loss, start loss, or transcript amplification) and causing LoF on the canonical transcript. Whole genomes and exomes were from patients with AD, healthy older controls, and a mix of subjects with other diagnoses including mild cognitive impairment, corticobasal degeneration, and other dementia not otherwise specified.

### Structural variant identification

ADSP release 3 (R3) individual-level VCF structural variant (SV) calls from software packages Manta, Smoove, and joint genotyping VCF from Biograph were downloaded from NIAGADS 8^th^ (Manta, Smoove) and 9^th^ releases (Biograph) (https://dss.niagads.org/datasets/ng00067/#data-releases). Insertions (SVTYPE INS), deletions (SVTYPE DEL), inversions (SVTYPE INV or intra-chromosomal SVTYPE BND with INFO/EVENT field), duplications (SVTYPE DUP), and translocations (SVTYPE TRA or inter-chromosomal SVTYPE BND with INFO/EVENT field) up to 20 kilobases in length that overlap *APOE* (hg38; chr19: 44905796-44909393) were identified via the following approach. Variants matching the above criteria on chromosome 19 with start position within the range (44905796, 44909393), as well as variants on chromosome 19 with start position within the range (44905796 – 20000, 44909393) and end position within the range (44905796, 44909393 + 20000), were isolated. Breakend calls not matching a variant type described above (i.e. SVTYPE BND calls without INFO/EVENT field) were excluded.

### Distribution of Tau Braak staging and neuritic plaques density in function of age-at-death in ε3/ε4

The uniform data set (UDS) obtained from the National Alzheimer’s Coordinating Center (NACC), December 2020 data freeze, was queried for individuals with *APOE* genotype, recorded age-at-death and neuropathological assessment available in NACC UDS (5,168 individuals). To assess where the W5*-ε3/ε4 carrier stands compared to other ε3/ε4 subjects, we subset this dataset to ε3/ε4 individuals who died after 60 years old, leading to 1,758 individuals with CERAD (Consortium to Establish a Registry for Alzheimer’s Disease) score of neuritic plaques density and 1,750 individuals with tau Braak staging. The density plots (**Figure 2**) were made with the *kdeplot* function of the *seaborn (*v.0.12) package in Python (v.3.7.7).

### DNA/RNA extraction from brain tissue

Genomic DNA and RNA were isolated from frozen post-mortem brain using the AllPrep DNA/RNA Mini Kit (Qiagen). Subject 6 nucleic acids were extracted from frontal lobe tissue and Subject 1 nucleic acids were extracted from cerebellum tissue. Nucleic acid concentrations were measured by NanoPhotometer (Implen), and DNA was stored at −20°C and RNA was stored at −80°C prior to use.

### Reverse transcriptase (RT) reaction

Total RNA (100 ng) was used for each 20 μL RT reaction, and cDNA synthesis was performed with random primers using the PrimeScript RT Reagent Kit (Takara Bio USA).

### DNA cloning

A primer pair AE-Ex1_F_pGL4-XhoI and AE-3’UTR_R_pGL4-Hind3 (**Supplementary Table 2**) was used to amplify full-length *APOE* cDNA that was reverse transcribed from Subject 1 RNA. The PCR profile consisted of 15 min at 95°C, 35 cycles of 20 sec at 95°C, 20 sec at 55°C, and 3 min at 72°C. The amplified fragment was inserted into pGL4.10[luc2] vector (Promega) that was cut with XhoI and HindIII using the In-Fusion® HD Cloning Kit (Takara Clontech). The In-Fusion ligation mix was transformed into Stellar™ Competent Cells (Takara Clontech). The cells were plated on LB Agar Plate containing 100 ug/ml ampicillin and was incubated at 37°C for 16 hours.

### Sanger sequencing

DNA was PCR amplified using the primers AE-Ex1_F_pGL4-XhoI and AE-3’UTR_R_pGL4-Hind3 (**Supplementary Table 2**). The PCR profile consisted of 15 min at 95°C, 35 cycles of 20 sec at 95°C, 20 sec at 55°C, and 3 min at 72°C. The PCR amplified DNAs were sequenced using the BigDye™ Terminator v3.1 Cycle Sequencing Kit (ThermoFisher Scientific). Primers Ch19_50103673_F and Ch19_50103049_R were used to obtain sequencing reads for SNP rs429358 and for the mutation located at rs777551553 on the *APOE* gene, respectively. The sequencing profile consisted of 1 min at 96°C, 35 cycles of 30 sec at 96°C, 10 sec at 55°C, and 4 min at 60°C.

### Long-read whole genome sequencing

Whole blood collected in EDTA and stored at −80C was used to extract genomic DNA using PureGene kit (Qiagen) for Subject 2. A sequencing library was prepared using Oxford Nanopore Rapid sequencing protocol (Oxford Nanopore Technologies, UK). The library was distributed over 3 R9 PromethION flow-cells and sequenced for a total of 24 hours on a PromethION48 sequencing device, achieving a total output of 116 gigabases of data. Fast5 files were base called using Guppy V6.4.2 Super Accurate(ONT) and aligned using MiniMap2^20^.

### Determination of mean AAO by *APOE* genotype

Genetic and phenotypic data from AD-related cohorts from the Alzheimer’s Disease Genetics Consortium (ADGC) and Alzheimer’s Disease Sequencing Project (ADSP) were processed as previously described^21^. This includes state-of-the-art quality control of *APOE* ε4 status and resolving of phenotypes and age information across duplicated samples. Unique, non-duplicate subjects (identified using identity-by-descent; Plink v1.9) that were non-Hispanic and of European ancestry (SNPWeights v2.1^22^) were retained to determine mean age-at-onset across *APOE* strata.

### Cell culture, transfection and monoclonal heterozygous p.W228* line generation

Human hepatocyte line HepG2 cells were grown in Dulbecco’s Modified Eagle Medium (DMEM) (Gibco) supplemented with 10% fetal bovine serum (FBS) (Gibco) and 1% Penicillin/Streptomycin (Gibco) at 37 ℃, 5% CO_2_. For prime editing, 150,000 cells were seeded one day prior to transfection at a density of 150,000 cells/well in 24-well plate. On the day of transfection, using 1 μL jetOPTIMUS (Polyplus) per well, following the manufacturer’s instructions. Cells were transfected with 750Lng of the pCMV-PE2 (Addgene #132775), 83Lng of the pegRNA and 83 ng of the nicking sgRNA per well. 72 hr post transfection, single cells were sorted into 96-well plates using BD FACSAria II SORP and cultured until confluency.

To assess prime editing, loci were amplified from isolated single clone genomic DNA samples via two rounds of PCR then deep sequenced. Briefly, the first round PCR (PCR1) amplified the genomic sequence of interest using primers containing Illumina forward and reverse adapters: NGS-ApoE-F: 5’ CCATCTCATCCCTGCGTGTCTCCCAAGCTGCGTAAGCGGCTCCTC 3’

NGS-ApoE-R: 5’ CCTCTCTATGGGCAGTCGGTGATGCACCTGCTCCTTCACCTCGTC 3’

The second round PCR step (PCR2) added unique i7 and i5 index combinations to both ends of the PCR1 product:

CL_AmpNGS_BC_F: 5’ AATGATACGGCGACCACCGAGATCTACAC[8nt-barcode]ACACTCTTTCCCTACACGACGCTCTTCCGATCT[0-11nt stager]CCATCTCATCCCTGCGTGTCTCC 3’

CL_AmpNGS_BC_R: 5’ CAAGCAGAAGACGGCATACGAGAT[[8nt-barcode]GTGACTGGAGTTCAGACGTGTGCTCTTCCGATCT[0-11nt stager]CCTCTCTATGGGCAGTCGGTGATg 3’

The amplified products were quantified with Qubit™ dsDNA HS Assay Kit (Thermo Fisher Scientific) and normalized by concentration, followed by sequencing using Illumina Miseq Reagent Kit v3 then analyzed with the CRISPResso2.

### ELISA Measurement of apoE protein level in prime-edited p.W228* cells

Monoclonal gene-edited cells that harbor both the wild-type and the p.W228* alleles at the *APOE* locus were maintained in culture for at least 48 hours. Afterwards, the cells were harvested and lysed with RIPA buffer (Cell Signaling Technology) to extract the total proteins. Lysate containing the protein extracts were then subjected to ELISA detection using human apoE ELISA kit (Mabtech) following manufacturer’s recommended protocols. Data were analyzed using Prism9.

## Acknowledgements

This work was supported by the following grants: RO1AG060747; R35AG072290; P30AG066515; P30AG066511; P30AG066509, as well as Merit Review Awards (BX000933 and BX004823) from the U.S. Department of Veterans Affairs Office of Research and Development Biomedical Laboratory Research Program, National Institutes of Health (P30AG066509, University of Washington Alzheimer’s Disease Research Center), U19 AG066567, Adult Changes in Thought study; T32 AG052354-06A1 UW AD Training grant, to D.D.C.), and the Nancy and Buster Alvord endowment (to C.D.K). This study was also supported by grants from NIH K99AG075238 (M.E.B.), P30AG066515 Development project (M.E.B.), the Alzheimer’s Association (AARF-20-683984, M.E.B.), and the European Union’s Horizon 2020 research and innovation program under the Marie Skłodowska-Curie (grant agreement No. 890650, Y.L.G.). The contents do not represent the views of the U.S. Department of Veterans Affairs or the United States Government.

ADGC is funded by NIA/NIH Grant U01 AG032984. The NACC database is funded by NIA/NIH Grant U24 AG072122. NACC data are contributed by the NIA-funded ADRCs: P30 AG062429 (PI James Brewer, MD, PhD), P30 AG066468 (PI Oscar Lopez, MD), P30 AG062421 (PI Bradley Hyman, MD, PhD), P30 AG066509 (PI Thomas Grabowski, MD), P30 AG066514 (PI Mary Sano, PhD), P30 AG066530 (PI Helena Chui, MD), P30 AG066507 (PI Marilyn Albert, PhD), P30 AG066444 (PI John Morris, MD), P30 AG066518 (PI Jeffrey Kaye, MD), P30 AG066512 (PI Thomas Wisniewski, MD), P30 AG066462 (PI Scott Small, MD), P30 AG072979 (PI David Wolk, MD), P30 AG072972 (PI Charles DeCarli, MD), P30 AG072976 (PI Andrew Saykin, PsyD), P30 AG072975 (PI David Bennett, MD), P30 AG072978 (PI Neil Kowall, MD), P30 AG072977 (PI Robert Vassar, PhD), P30 AG066519 (PI Frank LaFerla, PhD), P30 AG062677 (PI Ronald Petersen, MD, PhD), P30 AG079280 (PI Eric Reiman, MD), P30 AG062422 (PI Gil Rabinovici, MD), P30 AG066511 (PI Allan Levey, MD, PhD), P30 AG072946 (PI Linda Van Eldik, PhD), P30 AG062715 (PI Sanjay Asthana, MD, FRCP), P30 AG072973 (PI Russell Swerdlow, MD), P30 AG066506 (PI Todd Golde, MD, PhD), P30 AG066508 (PI Stephen Strittmatter, MD, PhD), P30 AG066515 (PI Victor Henderson, MD, MS), P30 AG072947 (PI Suzanne Craft, PhD), P30 AG072931 (PI Henry Paulson, MD, PhD), P30 AG066546 (PI Sudha Seshadri, MD), P20 AG068024 (PI Erik Roberson, MD, PhD), P20 AG068053 (PI Justin Miller, PhD), P20 AG068077 (PI Gary Rosenberg, MD), P20 AG068082 (PI Angela Jefferson, PhD), P30 AG072958 (PI Heather Whitson, MD), P30 AG072959 (PI James Leverenz, MD).

Data for this study were prepared, archived, and distributed by the National Institute on Aging Alzheimer’s Disease Data Storage Site (NIAGADS) at the University of Pennsylvania (U24-AG041689), funded by the National Institute on Aging.

The Alzheimer’s Disease Sequencing Project (ADSP) is comprised of two Alzheimer’s Disease (AD) genetics consortia and three National Human Genome Research Institute (NHGRI) funded Large Scale Sequencing and Analysis Centers (LSAC). The two AD genetics consortia are the Alzheimer’s Disease Genetics Consortium (ADGC) funded by NIA (U01 AG032984), and the Cohorts for Heart and Aging Research in Genomic Epidemiology (CHARGE) funded by NIA (R01 AG033193), the National Heart, Lung, and Blood Institute (NHLBI), other National Institute of Health (NIH) institutes and other foreign governmental and non-governmental organizations. The Discovery Phase analysis of sequence data is supported through UF1AG047133 (to Drs. Schellenberg, Farrer, Pericak-Vance, Mayeux, and Haines); U01AG049505 to Dr. Seshadri; U01AG049506 to Dr. Boerwinkle; U01AG049507 to Dr. Wijsman; and U01AG049508 to Dr. Goate and the Discovery Extension Phase analysis is supported through U01AG052411 to Dr. Goate, U01AG052410 to Dr. Pericak-Vance and U01 AG052409 to Drs. Seshadri and Fornage. Sequencing for the Follow Up Study (FUS) is supported through U01AG057659 (to Drs. PericakVance, Mayeux, and Vardarajan) and U01AG062943 (to Drs. Pericak-Vance and Mayeux). Data generation and harmonization in the Follow-up Phase is supported by U54AG052427 (to Drs. Schellenberg and Wang). The FUS Phase analysis of sequence data is supported through U01AG058589 (to Drs. Destefano, Boerwinkle, De Jager, Fornage, Seshadri, and Wijsman), U01AG058654 (to Drs. Haines, Bush, Farrer, Martin, and Pericak-Vance), U01AG058635 (to Dr. Goate), RF1AG058066 (to Drs. Haines, Pericak-Vance, and Scott), RF1AG057519 (to Drs. Farrer and Jun), R01AG048927 (to Dr. Farrer), and RF1AG054074 (to Drs. Pericak-Vance and Beecham).

The ADGC cohorts include: Adult Changes in Thought (ACT) (U01 AG006781, U19 AG066567), the Alzheimer’s Disease Research Centers (ADRC) (P30 AG062429, P30 AG066468, P30 AG062421, P30 AG066509, P30 AG066514, P30 AG066530, P30 AG066507, P30 AG066444, P30 AG066518, P30 AG066512, P30 AG066462, P30 AG072979, P30 AG072972, P30 AG072976, P30 AG072975, P30 AG072978, P30 AG072977, P30 AG066519, P30 AG062677, P30 AG079280, P30 AG062422, P30 AG066511, P30 AG072946, P30 AG062715, P30 AG072973, P30 AG066506, P30 AG066508, P30 AG066515, P30 AG072947, P30 AG072931, P30 AG066546, P20 AG068024, P20 AG068053, P20 AG068077, P20 AG068082, P30 AG072958, P30 AG072959), the Chicago Health and Aging Project (CHAP) (R01 AG11101, RC4 AG039085, K23 AG030944), Indiana Memory and Aging Study (IMAS) (R01 AG019771), Indianapolis Ibadan (R01 AG009956, P30 AG010133), the Memory and Aging Project (MAP) (R01 AG17917), Mayo Clinic (MAYO) (R01 AG032990, U01 AG046139, R01 NS080820, RF1 AG051504, P50 AG016574), Mayo Parkinson’s Disease controls (NS039764, NS071674, 5RC2HG005605), University of Miami (R01 AG027944, R01 AG028786, R01 AG019085, IIRG09133827, A2011048), the Multi-Institutional Research in Alzheimer’s Genetic Epidemiology Study (MIRAGE) (R01 AG09029, R01 AG025259), the National Centralized Repository for Alzheimer’s Disease and Related Dementias (NCRAD) (U24 AG021886), the National Institute on Aging Late Onset Alzheimer’s Disease Family Study (NIA-LOAD) (U24 AG056270), the Religious Orders Study (ROS) (P30 AG10161, R01 AG15819), the Texas Alzheimer’s Research and Care Consortium (TARCC) (funded by the Darrell K Royal Texas Alzheimer’s Initiative), Vanderbilt University/Case Western Reserve University (VAN/CWRU) (R01 AG019757, R01 AG021547, R01 AG027944, R01 AG028786, P01 NS026630, and Alzheimer’s Association), the Washington Heights-Inwood Columbia Aging Project (WHICAP) (RF1 AG054023), the University of Washington Families (VA Research Merit Grant, NIA: P50AG005136, R01AG041797, NINDS: R01NS069719), the Columbia University Hispanic Estudio Familiar de Influencia Genetica de Alzheimer (EFIGA) (RF1 AG015473), the University of Toronto (UT) (funded by Wellcome Trust, Medical Research Council, Canadian Institutes of Health Research), and Genetic Differences (GD) (R01 AG007584). The CHARGE cohorts are supported in part by National Heart, Lung, and Blood Institute (NHLBI) infrastructure grant HL105756 (Psaty), RC2HL102419 (Boerwinkle) and the neurology working group is supported by the National Institute on Aging (NIA) R01 grant AG033193.

The CHARGE cohorts participating in the ADSP include the following: Austrian Stroke Prevention Study (ASPS), ASPS-Family study, and the Prospective Dementia Registry-Austria (ASPS/PRODEM-Aus), the Atherosclerosis Risk in Communities (ARIC) Study, the Cardiovascular Health Study (CHS), the Erasmus Rucphen Family Study (ERF), the Framingham Heart Study (FHS), and the Rotterdam Study (RS). ASPS is funded by the Austrian Science Fond (FWF) grant number P20545-P05 and P13180 and the Medical University of Graz. The ASPS-Fam is funded by the Austrian Science Fund (FWF) project I904), the EU Joint Programme – Neurodegenerative Disease Research (JPND) in frame of the BRIDGET project (Austria, Ministry of Science) and the Medical University of Graz and the Steiermärkische Krankenanstalten Gesellschaft. PRODEM-Austria is supported by the Austrian Research Promotion agency (FFG) (Project No. 827462) and by the Austrian National Bank (Anniversary Fund, project 15435. ARIC research is carried out as a collaborative study supported by NHLBI contracts (HHSN268201100005C, HHSN268201100006C, HHSN268201100007C, HHSN268201100008C, HHSN268201100009C, HHSN268201100010C, HHSN268201100011C, and HHSN268201100012C). Neurocognitive data in ARIC is collected by U01 2U01HL096812, 2U01HL096814, 2U01HL096899, 2U01HL096902, 2U01HL096917 from the NIH (NHLBI, NINDS, NIA and NIDCD), and with previous brain MRI examinations funded by R01-HL70825 from the NHLBI. CHS research was supported by contracts HHSN268201200036C, HHSN268200800007C, N01HC55222, N01HC85079, N01HC85080, N01HC85081, N01HC85082, N01HC85083, N01HC85086, and grants U01HL080295 and U01HL130114 from the NHLBI with additional contribution from the National Institute of Neurological Disorders and Stroke (NINDS). Additional support was provided by R01AG023629, R01AG15928, and R01AG20098 from the NIA. FHS research is supported by NHLBI contracts N01-HC-25195 and HHSN268201500001I. This study was also supported by additional grants from the NIA (R01s AG054076, AG049607 and AG033040 and NINDS (R01 NS017950). The ERF study as a part of EUROSPAN (European Special Populations Research Network) was supported by European Commission FP6 STRP grant number 018947 (LSHG-CT-2006-01947) and also received funding from the European Community’s Seventh Framework Programme (FP7/2007-2013)/grant agreement HEALTH-F4-2007-201413 by the European Commission under the programme “Quality of Life and Management of the Living Resources” of 5th Framework Programme (no. QLG2-CT-2002-01254). High-throughput analysis of the ERF data was supported by a joint grant from the Netherlands Organization for Scientific Research and the Russian Foundation for Basic Research (NWO-RFBR 047.017.043). The Rotterdam Study is funded by Erasmus Medical Center and Erasmus University, Rotterdam, the Netherlands Organization for Health Research and Development (ZonMw), the Research Institute for Diseases in the Elderly (RIDE), the Ministry of Education, Culture and Science, the Ministry for Health, Welfare and Sports, the European Commission (DG XII), and the municipality of Rotterdam. Genetic data sets are also supported by the Netherlands Organization of Scientific Research NWO Investments (175.010.2005.011, 911-03-012), the Genetic Laboratory of the Department of Internal Medicine, Erasmus MC, the Research Institute for Diseases in the Elderly (014-93-015; RIDE2), and the Netherlands Genomics Initiative (NGI)/Netherlands Organization for Scientific Research (NWO) Netherlands Consortium for Healthy Aging (NCHA), project 050-060-810. All studies are grateful to their participants, faculty and staff. The content of these manuscripts is solely the responsibility of the authors and does not necessarily represent the official views of the National Institutes of Health or the U.S. Department of Health and Human Services.

The FUS cohorts include: the Alzheimer’s Disease Research Centers (ADRC) (P30 AG062429, P30 AG066468, P30 AG062421, P30 AG066509, P30 AG066514, P30 AG066530, P30 AG066507, P30 AG066444, P30 AG066518, P30 AG066512, P30 AG066462, P30 AG072979, P30 AG072972, P30 AG072976, P30 AG072975, P30 AG072978, P30 AG072977, P30 AG066519, P30 AG062677, P30 AG079280, P30 AG062422, P30 AG066511, P30 AG072946, P30 AG062715, P30 AG072973, P30 AG066506, P30 AG066508, P30 AG066515, P30 AG072947, P30 AG072931, P30 AG066546, P20 AG068024, P20 AG068053, P20 AG068077, P20 AG068082, P30 AG072958, P30 AG072959), Alzheimer’s Disease Neuroimaging Initiative (ADNI) (U19AG024904), Amish Protective Variant Study (RF1AG058066), Cache County Study (R01AG11380, R01AG031272, R01AG21136, RF1AG054052), Case Western Reserve University Brain Bank (CWRUBB) (P50AG008012), Case Western Reserve University Rapid Decline (CWRURD) (RF1AG058267, NU38CK000480), CubanAmerican Alzheimer’s Disease Initiative (CuAADI) (3U01AG052410), Estudio Familiar de Influencia Genetica en Alzheimer (EFIGA) (5R37AG015473, RF1AG015473, R56AG051876), Genetic and Environmental Risk Factors for Alzheimer Disease Among African Americans Study (GenerAAtions) (2R01AG09029, R01AG025259, 2R01AG048927), Gwangju Alzheimer and Related Dementias Study (GARD) (U01AG062602), Hillblom Aging Network (2014-A-004-NET, R01AG032289, R01AG048234), Hussman Institute for Human Genomics Brain Bank (HIHGBB) (R01AG027944, Alzheimer’s Association “Identification of Rare Variants in Alzheimer Disease”), Ibadan Study of Aging (IBADAN) (5R01AG009956), Longevity Genes Project (LGP) and LonGenity (R01AG042188, R01AG044829, R01AG046949, R01AG057909, R01AG061155, P30AG038072), Mexican Health and Aging Study (MHAS) (R01AG018016), Multi-Institutional Research in Alzheimer’s Genetic Epidemiology (MIRAGE) (2R01AG09029, R01AG025259, 2R01AG048927), Northern Manhattan Study (NOMAS) (R01NS29993), Peru Alzheimer’s Disease Initiative (PeADI) (RF1AG054074), Puerto Rican 1066 (PR1066) (Wellcome Trust (GR066133/GR080002), European Research Council (340755)), Puerto Rican Alzheimer Disease Initiative (PRADI) (RF1AG054074), Reasons for Geographic and Racial Differences in Stroke (REGARDS) (U01NS041588), Research in African American Alzheimer Disease Initiative (REAAADI) (U01AG052410), the Religious Orders Study (ROS) (P30 AG10161, P30 AG72975, R01 AG15819, R01 AG42210), the RUSH Memory and Aging Project (MAP) (R01 AG017917, R01 AG42210Stanford Extreme Phenotypes in AD (R01AG060747), University of Miami Brain Endowment Bank (MBB), University of Miami/Case Western/North Carolina A&T African American (UM/CASE/NCAT) (U01AG052410, R01AG028786), and Wisconsin Registry for Alzheimer’s Prevention (WRAP) (R01AG027161 and R01AG054047).

The four LSACs are: the Human Genome Sequencing Center at the Baylor College of Medicine (U54 HG003273), the Broad Institute Genome Center (U54HG003067), The American Genome Center at the Uniformed Services University of the Health Sciences (U01AG057659), and the Washington University Genome Institute (U54HG003079). Genotyping and sequencing for the ADSP FUS is also conducted at John P. Hussman Institute for Human Genomics (HIHG) Center for Genome Technology (CGT).

Biological samples and associated phenotypic data used in primary data analyses were stored at Study Investigators institutions, and at the National Centralized Repository for Alzheimer’s Disease and Related Dementias (NCRAD, U24AG021886) at Indiana University funded by NIA. Associated Phenotypic Data used in primary and secondary data analyses were provided by Study Investigators, the NIA funded Alzheimer’s Disease Centers (ADCs), and the National Alzheimer’s Coordinating Center (NACC, U24AG072122) and the National Institute on Aging Genetics of Alzheimer’s Disease Data Storage Site (NIAGADS, U24AG041689) at the University of Pennsylvania, funded by NIA. Harmonized phenotypes were provided by the ADSP Phenotype Harmonization Consortium (ADSP-PHC), funded by NIA (U24 AG074855, U01 AG068057 and R01 AG059716) and Ultrascale Machine Learning to Empower Discovery in Alzheimer’s Disease Biobanks (AI4AD, U01 AG068057). This research was supported in part by the Intramural Research Program of the National Institutes of health, National Library of Medicine. Contributors to the Genetic Analysis Data included Study Investigators on projects that were individually funded by NIA, and other NIH institutes, and by private U.S. organizations, or foreign governmental or nongovernmental organizations.

The Alzheimer’s Disease Genetics Consortium (ADGC) supported sample preparation, sequencing and data processing through NIA grant U01AG032984. Sequencing data generation and harmonization is supported by the Genome Center for Alzheimer’s Disease, U54AG052427, and data sharing is supported by NIAGADS, U24AG041689. Samples from the National Centralized Repository for Alzheimer’s Disease and Related Dementias (NCRAD), which receives government support under a cooperative agreement grant (U24 AG021886) awarded by the National Institute on Aging (NIA), were used in this study. We thank contributors who collected samples used in this study, as well as patients and their families, whose help and participation made this work possible.

