## Supplementary Figures for "*APOE* loss-of-function variants: Compatible with longevity and associated with resistance to Alzheimer’s Disease pathology"

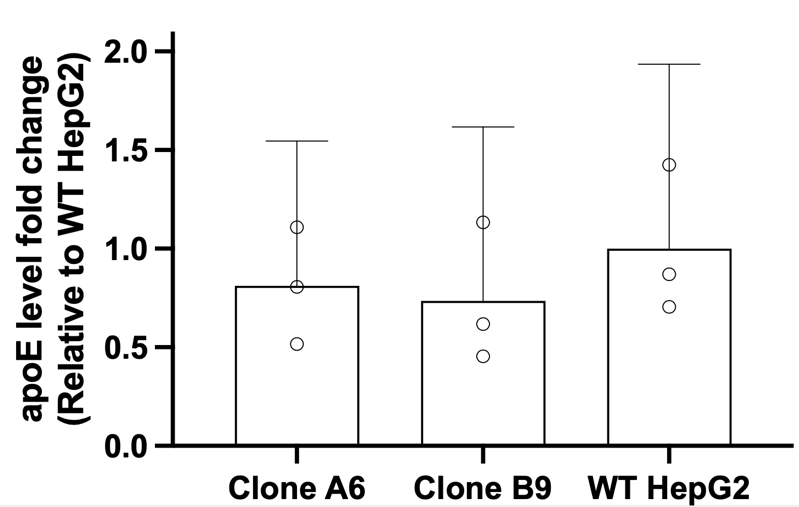

**Figure S1. Assessing APOE protein level in hepatocytes carrying variant p.W228*.** There was no significant difference in APOE protein levels measured by ELISA between WT hepatocytes and two hepatocyte clonal lines modified by prime editing to be heterozygous for p.W228*.

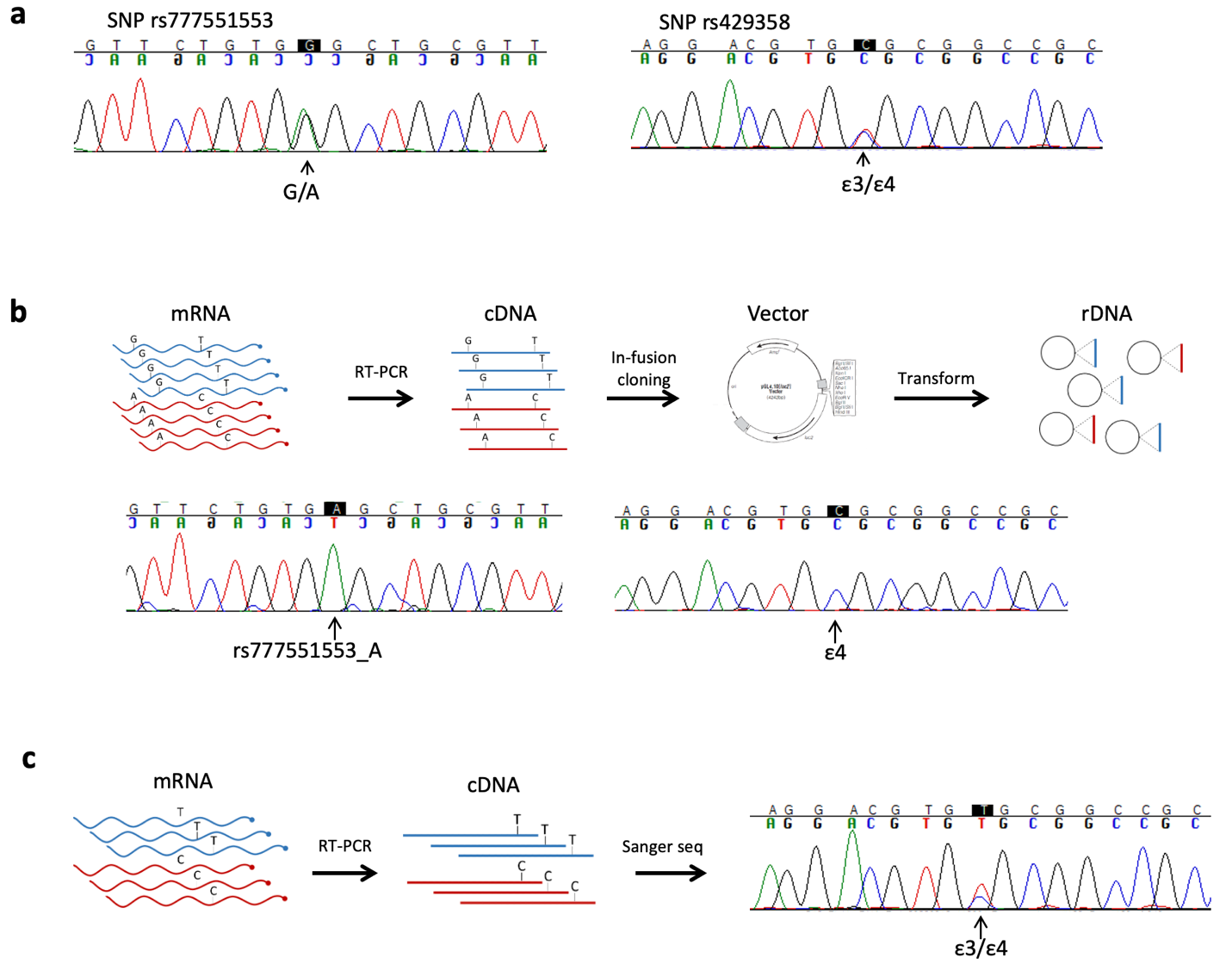

**Figure S2. Subject 1 genotyping, rs777551553 phasing, and expression analysis.** (a) Sanger sequencing confirms that Subject 1 is *APOE* ε3/ε4 and heterozygous for rs777551553_A. (b) Cloning of reverse transcribed mRNA demonstrates that rs777551553_A is in phase with ε4. (c) Both ε3 and ε4 are expressed in post-mortem cerebellar tissue.

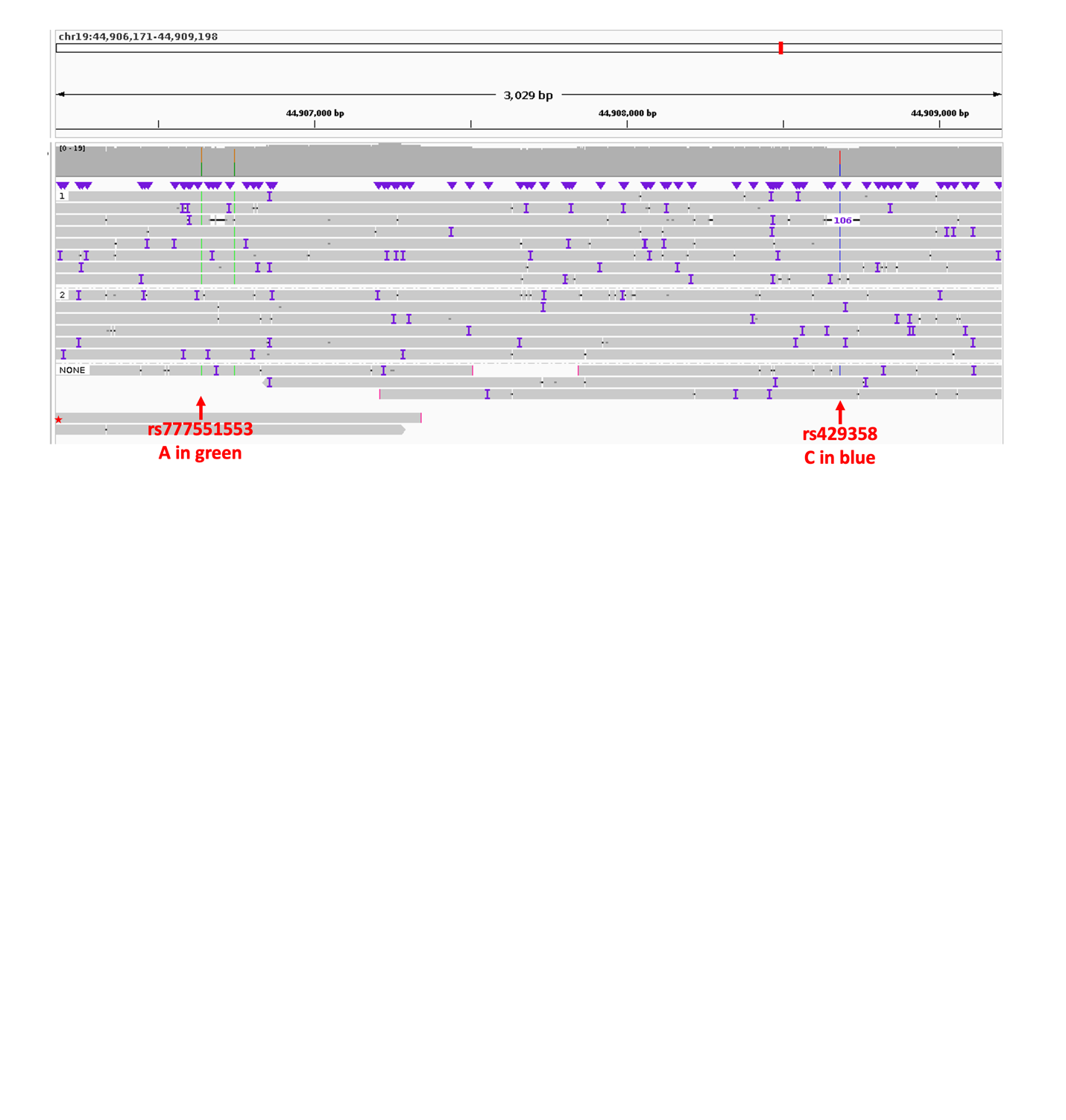

**Figure S3. In Subject 2, confirmation of rs777551553_A variant call and phasing.** 12 reads cover both SNP positions in LRS; 5 reads carry both rs777551553_A (stop gain) and rs429358_C (ε4), and 6 reads carry both rs777551553_G (WT) and rs429358_T (ε3). This is consistent with the stop gain and the ε4 on the same chromosome. WES also confirmed that Subject 2 is ε3/ε4 (total read count 18; T = 9; C = 9) and is heterozygous for rs777551553_A (total read count 39; G = 20; A = 19) (data not shown).

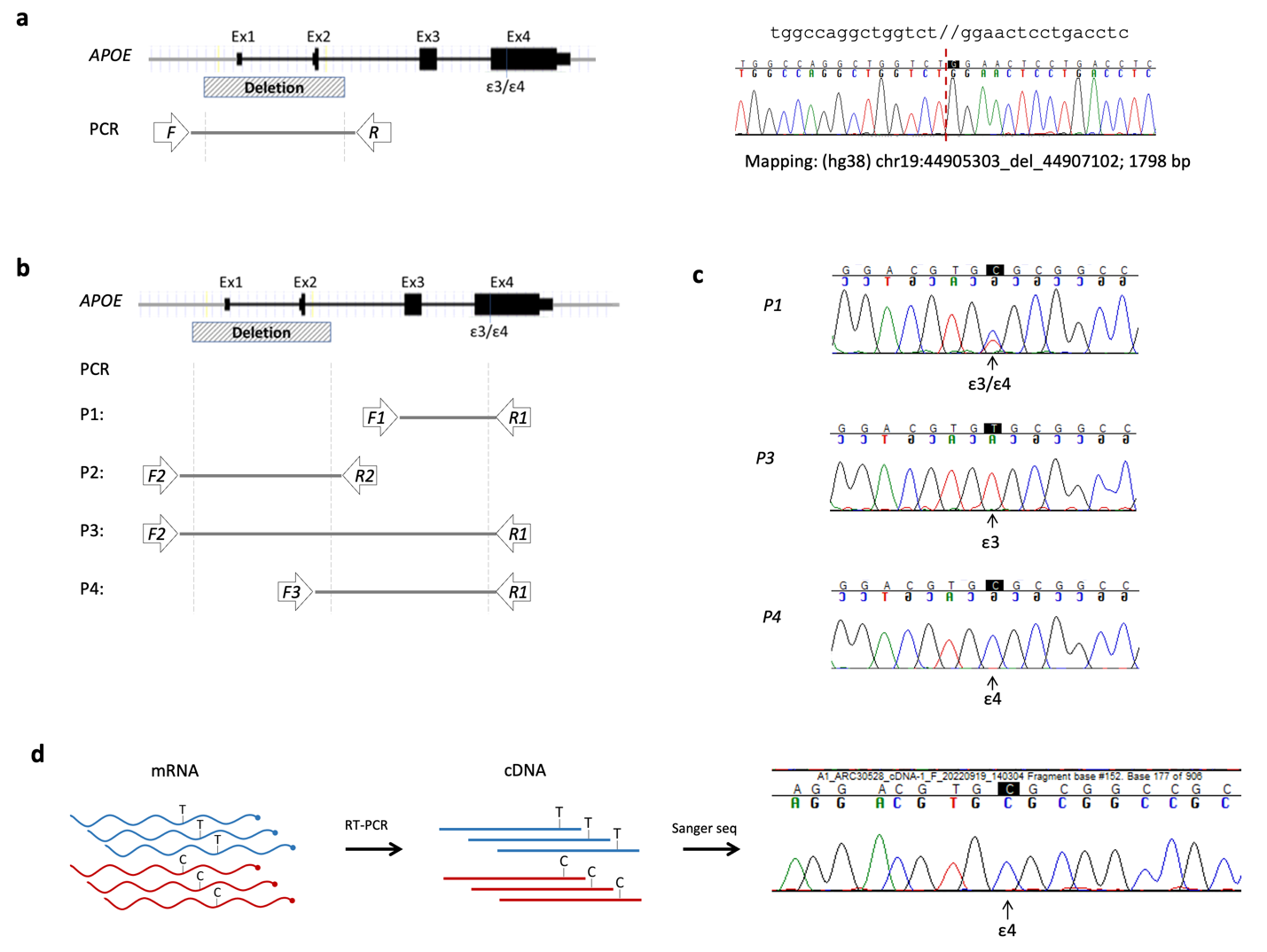

**Figure S4. Subject 6 *APOE* genotyping, *APOE* deletion phasing and expression analysis.** (a) Sanger sequencing was used to confirm the start and end positions of the deletion. (b) Primer sets were designed to phase the 1798 base pair deletion with ε3/4. (c) Sanger sequencing of PCR products establishes that both ε3 and ε4 alleles are present in post-mortem brain tissue and that the deletion is in phase with ε3. (d) Sanger sequencing of reverse transcribed mRNA from post-mortem frontal cortex tissue establishes that only the ε4 allele is expressed in Subject 6.
